# Changing patterns of cigarette and ENDS transitions in the USA: a multistate transition analysis of adults in the PATH Study in 2017–2019 vs 2019–2021

**DOI:** 10.1101/2023.10.20.23297320

**Authors:** Andrew F. Brouwer, Jihyoun Jeon, Evelyn Jimenez-Mendoza, Stephanie R. Land, Theodore R. Holford, Abigail S. Friedman, Jamie Tam, Ritesh Mistry, David T. Levy, Rafael Meza

## Abstract

**Introduction:** The use of cigarettes and electronic nicotine delivery system (ENDS) has likely changed since 2019 with the rise of pods and disposables, the outbreak of lung injuries related to vaping THC, flavor bans, and the COVID pandemic. We analyzed patterns of initiation, cessation, and transitions between cigarettes, ENDS, and dual use before and after 2019.

**Methods:** Using the Population Assessment of Tobacco and Health (PATH) Study, we applied a multistate transition model to 28,061 adults in Waves 4–5 (2017–19) and 24,751 adults in Waves 5–6 (2019–21), estimating transition rates for initiation, cessation, and switching products for each period overall and by age group.

**Results:** Cigarette initiation among adults who never used either product decreased from 2017–19 to 2019–21, but ENDS initiation did not significantly change. Persistence of ENDS-only use remained high, with 75–80% still using ENDS only after 1 year. Cigarette-only use transitions remained similar, with about 88% remaining, 7% transitioning to non-current use, and 5% transitioning to dual or ENDS-only use. In contrast, dual use to ENDS-only transitions increased from 9.5% (95%CI: 7.3–11.7%) to 20.1% (95%CI: 17.5–22.7%) per year from 2017–19 to 2019–21, decreasing the persistence of dual use. The dual use to cigarette-only transition remained at about 25%. These changes were qualitatively similar across adult age groups, though adults ages 18–24 years exhibited the highest probability of switching from cigarette-only use to dual use and from dual use to ENDS-only use.

**Conclusions:** Persistence of ENDS use among adults remained high in 2019–21, but a larger fraction of dual users transitioned to ENDS-only use compared to 2017–19. Because the fraction of cigarette-only users switching to dual use remained low, the public health implications of the increased dual use to ENDS-only transition are minimal.

## Introduction

The landscape of tobacco and nicotine products in the US and other countries has evolved quickly over the past decade. Electronic nicotine delivery systems (ENDS), including e-cigarettes, have transformed from cig-a-likes, refillables, and tank systems—which initially used freebase nicotine that was unpalatable at higher nicotine concentrations—to pod mods and disposables that use nicotine salts, often perceived as less harsh.^1–5^ As ENDS products have changed, so too have patterns of transitions between ENDS and cigarette products.^6, 7^ Moreover, recent events and policies, including the lung injury outbreak in 2019,^8^ the COVID-19 pandemic, and the increasing restriction of ENDS flavors,^9^ may have further shifted patterns of use and transition between ENDS and cigarettes.

Understanding product transitions is key to determining the likely public health benefit or harm of ENDS.^10–12^ If ENDS promote cigarette cessation, reduced smoking, or divert those who would have otherwise smoked, there would be a benefit to public health.^13–16^ However, there remain concerns about youth initiation, particularly as many flavors are targeted to youth,^17, 18^ as well as concerns that ENDS may interfere with long-term cigarette cessation because of continued or enhanced nicotine addiction.^19–21^ Many of the clinical trials that suggested ENDS can facilitate cigarette cessation used early generation products that bear little resemblance to products more recently on the market,^13, 14^ and the real-world effectiveness may differ from the efficacy measured in clinical trials for many reasons. Hence, continued analysis of how actual transition rates change over time is important for tracking real-world associations between ENDS and other product use and providing the information necessary to project future public health outcomes.

In previous work, we analyzed transitions in the Population Assessment of Tobacco and Health (PATH) Study Waves 1–5 (2013–19), a nationally representative longitudinal survey of the US, using a multistate transition model.^6, 7^ Multistate transition models estimate the transition rates that underly observed transitions between product.^22–31^ In this analysis, we extend our models to compare transitions observed in PATH in 2017–19 to 2019–21.

## Methods

### Data

The Population Assessment of Tobacco and Health (PATH) Study is a nationally representative longitudinal cohort study of tobacco and nicotine product use behaviors in the US among the civilian, noninstitutionalized adult population. Our analysis compared 28,061 adults in the Wave 4 cohort in Waves 4–5 (Dec 2016 to Nov 2019, abbreviated as 2017–19) and 24,751 adults in the Wave 4 cohort in Waves 5–6 (Dec 2018 to Nov 2021, abbreviated as 2019–21). Although this analysis primarily focuses on the differences between 2017–19 and 2019–21, we include previous results in 2015–17 in the supplementary material for comparison purposes (Figures S1–S3).^7^

Follow-up time for participants was approximately two years between Waves 4 and 5 and between Waves 5 and 6, and follow-up time was treated as exactly two years in the model. The multistate transition framework explicitly accounts for the time between observations and for potential unobserved transitions, so one-year transition probabilities can be estimated from two-year data. We categorized participants as ages 18–24, 25–34, or 35–90 years. This set of groups was chosen because our transition estimates were not well-powered for participants ages 55–90 alone in 2019–21.

Each participant’s product use was categorized as never established use (of either product), non-current use, cigarette-only use, ENDS-only use, or dual use of cigarettes and ENDS, as in previous work.^6, 7^ Established cigarette use was defined as having smoked at least 100 cigarettes in one’s lifetime, and established ENDS use was defined as ever “fairly regularly” using ENDS. Current use of cigarettes and ENDS was defined as any past 30-day use by an established use of that product, and current dual use was defined as current use of both cigarettes and ENDS by established users of both products. Non-current use was defined as no past 30-day use of either product by a participant who had established use of one or both products. Characteristics of the populations are given in the supplementary material (Table S1). This analysis was not regulated as human subjects research (University of Michigan Institutional Review Board HUM00162265).

### Transition modeling

We applied our multistate transition model to analyze the underlying transition hazard rates between product use for adults overall and in each of the three age categories (18–24, 25–34, or 35–90 years) in each of the two time periods (2017–19, 2019–21). Multistate transition models are finite-state, stochastic process models that assume that transition hazard rates depend only on the current state and not on past states or transition history. Technical details of the multistate transition model are provided in the Supplemental Material. Using discrete-time observations, the model estimated instantaneous risk of transition from one state to another, i.e., transition hazard rates, which collectively define the probability of transitioning from one state to any other at a future time. We incorporated Wave 4 longitudinal survey weights into the model. Using the estimated transition rates, we estimated the 1-year transition probabilities for each group for the two time periods separately. Additionally, two-sided p-values for whether transition rates were significantly different between periods were calculated as described in the Supplemental Material using the replicate weights provided by PATH. Multistate transition models were estimated using the `wmsm` function^6^ in R (publicly available at https://tcors.umich.edu/Resources_Research.php), which is an extension of the `msm` function^32^ modified to incorporate participant weights.

## Results

Over 2019–21, the average weighted prevalence of never established use was 57.7%, the prevalence of non-current use was 23.7%, the prevalence of cigarette-only use was 14.2%, the prevalence of ENDS-only use was 2.5%, and the prevalence of dual cigarette and ENDS use was 1.9%.

### Transition hazard rates

There were three transitions that statistically significantly changed between 2017–19 and 2019–21 (Figure 1). The transition rate of cigarette initiation declined for adults in 2019–21 (p=0.02). The transition rate from ENDS-only use to non-current use increased in 2019–21 (p=0.005). The dual use to ENDS-only use transition also increased in 2019–21 rate (p<0.001).

**Figure 1:**
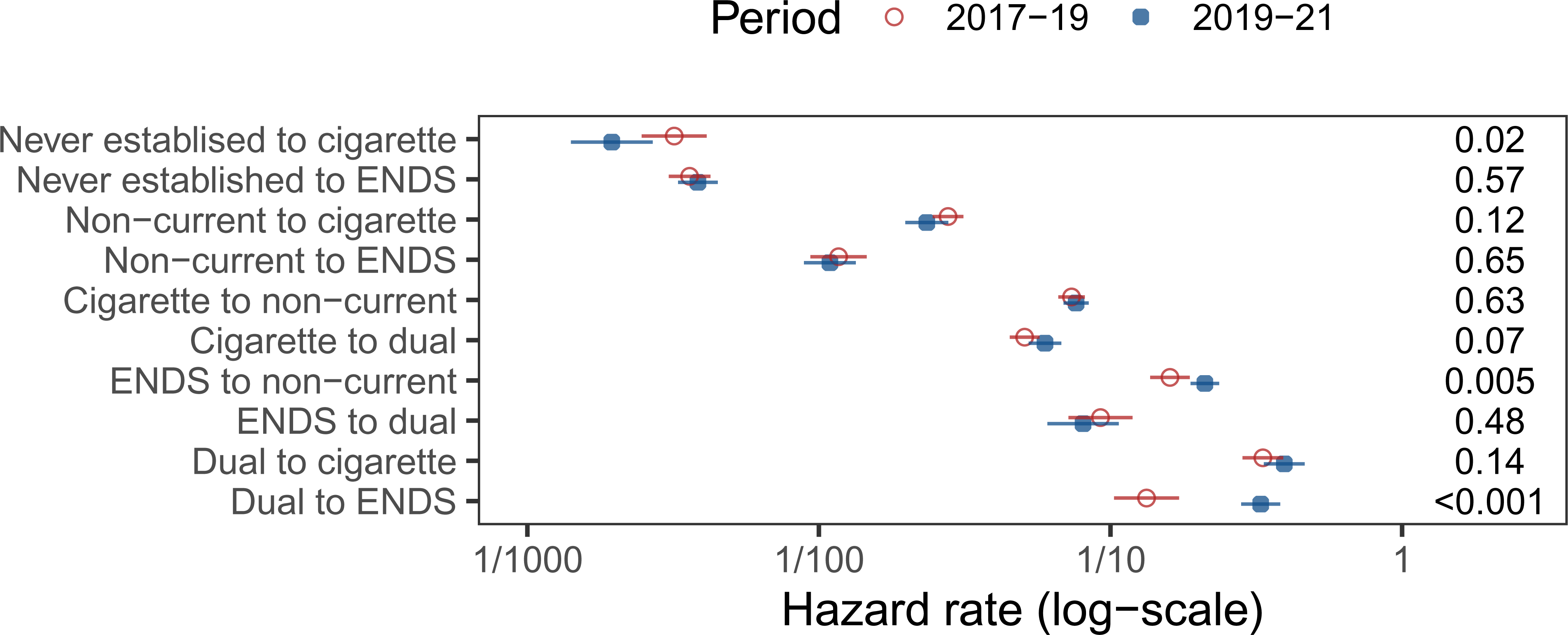
Transition hazard rates among adults in 2017–19 (Waves 4–5) and 2019–21 (Waves 5–6). A version of this figure including previous results from 2015–17 (Waves 2–4) is given in Figure S1.

### Transition probabilities

The changing transition rates resulted in changes in the 1-year transition probabilities (Figure 2). The transition patterns for those with never established use, non-current use, and cigarette only use remained largely similar between the two periods, with the large majority remaining in the same use state (never established use persistence: 98.8% (95% CI: 98.6–98.9%) and 98.6% (95% CI: 98.4–98.8%) in the two periods; non-current use persistence: 96.3% (95%CI: 95.9–96.7%) and 96.8% (95%CI: 96.4– 97.2%); and cigarette-only use: 88.2% (95%CI: 87.3–89.9%) and 88.2% (95%CI: 87.4–89.0%)). ENDS uptake among adults who only use cigarettes remained low, with 3.8% (95%CI: 3.4–4.2%) and 4.0% (95%CI: 3.5–4.4%) transitioning.

**Figure 2:**
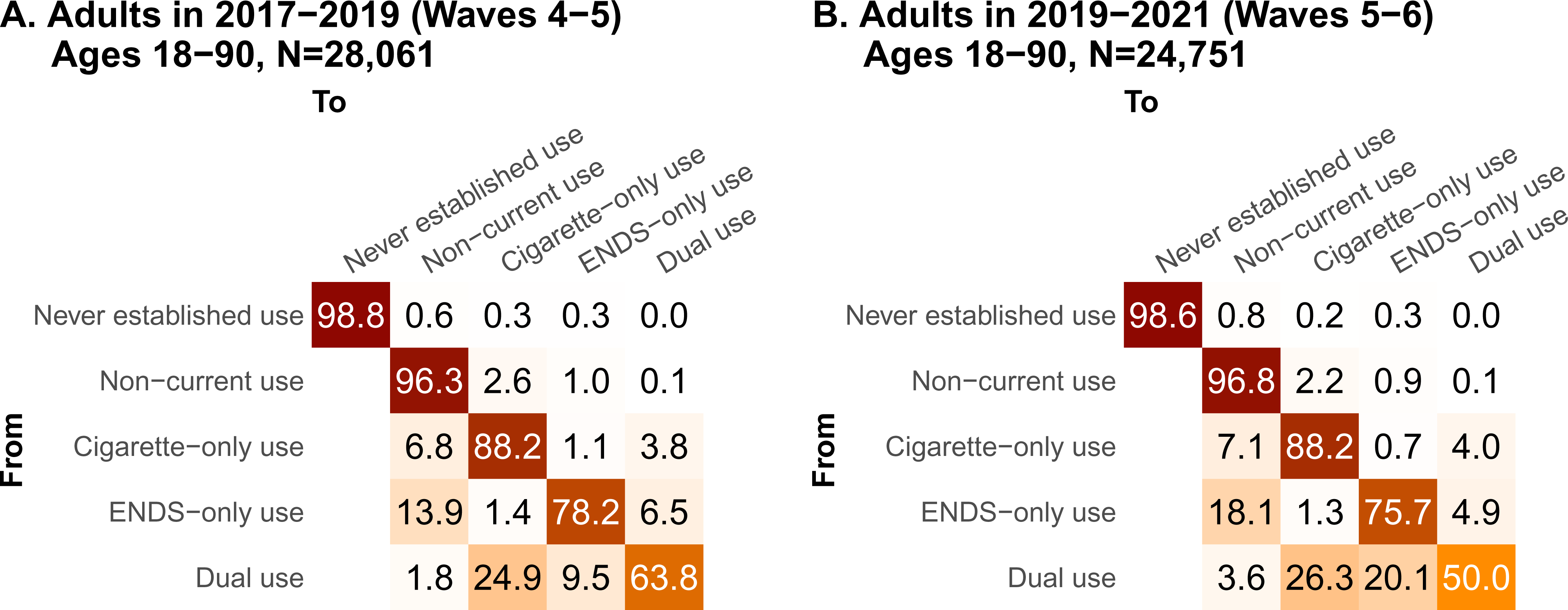
One-year transition probabilities for adults in A. 2017–19 (Waves 4–5), and B. 2019–21 (Waves 5–6). Confidence intervals, including previous results from 2015–17 (Waves 2–4), are shown in Figure S2. The transitions weighted by each use state’s prevalence are given in Figure S4.

The ENDS cessation probability increased slightly from 13.9% (95%CI: 11.9–16.0%) to 18.1% (95%CI: 16.2–19.9%). The transition patterns for those who dual used both products changed substantially over this period, driven by the increase in cigarette cessation. Specifically, from 2017–19 to 2019–21, the transition probability from dual to cigarette-only use did not significantly change, but the transition probability from dual to ENDS-only use nearly doubled, increasing from 9.5% (95%CI: 7.3–11.7%) to 20.1% (95%CI: 17.5–22.7%). This increase was at the expense of the persistence of dual use, which decreased from 63.8% (95%CI: 59.9–67.6%) to 50.0% (95%CI: 46.1–54.0%) between the periods. There was no analogous increase in the cigarette-only to non-current use transition, which remained low at 6.8% (95%CI 6.2–7.5%) and 7.1% (6.4–7.7%) in the two periods.

Considering the age-stratified transition probabilities (Figure 3), most of the changes in transitions for those using ENDS only or dual using both products were qualitatively similar across the age groups. The persistence of e-cigarette-only use remained similar in 2017–19 and 2019–21, decreasing somewhat for ages 25–34 only. Persistence of dual use decreased in 2019–21 for all three groups, accompanied by an increase in the transition probability of dual to ENDS-only.

**Figure 3:**
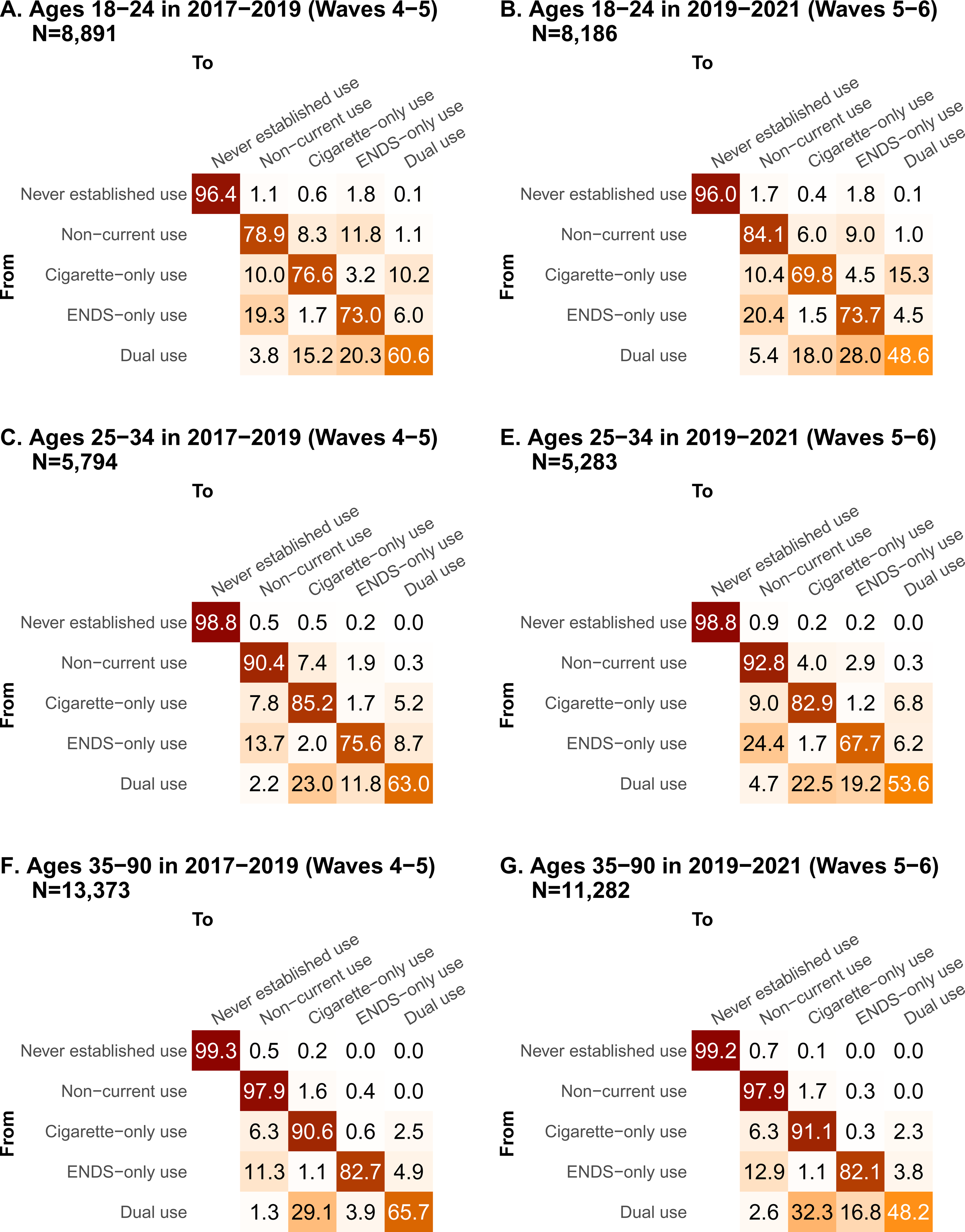
One-year transition probabilities for adults ages 18–24 (A–C), ages 25–34 (D–F), and ages 35– 90 (G–I). Each of the three plots compares periods 2015–17 (Waves 2–4), 2017–19 (Waves 4–5), and 2019–21 (Waves 5–6). Confidence intervals, including previous results from 2015–17 (Waves 2–4), are shown in Figure S3.

In 2019–21, young adults ages 18-24 were more likely to transition from cigarette-only use to dual use 15.3% (95%CI: 9.6–21.0%) than those ages 25–34 (6.8% (95%CI: 5.4–8.2%)) or ages 35–90 (2.3% (95%CI: 1.9–2.7%)). Additionally, while the dual use to ENDS-only transition increased for all age groups, it was higher for young adults (28.0% (95%CI: 21.6–34.4%) than for the other two ages groups (19.2% (95%CI: 14.2–24.1%) and 16.8% (95%CI: 13.0–20.7%), respectively).

## Discussion

Changing marketplaces, policy environments, public health crises, and evolving attitudes and perceptions of products have made it difficult to predict short-term patterns of use of cigarettes and ENDS, let alone their longer-term impacts on public health. In previous work, we had identified dramatic increases in the persistence of ENDS-only and dual use of cigarettes and ENDS around the time of the marketplace shift to pod mod products, including JUUL.^7^ Here, we found that many of the transition rates that significantly changed from 2015–17 to 2017–19 did not significantly change again in 2017–19 to 2019–21 (Figure S1), including the non-current to cigarette-only use, cigarette-only to non-current use, ENDS-only to dual use, and dual to cigarette-only use transitions. We did find three significant changes in transition rates from 2017–19 to 2019–21: a decline in never-established to cigarette-only use, an increase in ENDS-only to non-current use, and an increase in dual to ENDS-only use. From the perspective of reducing cigarette use, these changes appear to be beneficial, but the big picture underscores the overall public health challenge (Figure S4), with high persistence of cigarette, ENDS, and dual use. In particular, because dual use remained uncommon (1.9% among adults overall) and ENDS uptake among adults who smoke cigarettes remained low (<5% switching to dual or ENDS-only use in one year), particularly for those ages 25 and older, the population public health impact of the increase in the dual to ENDS-only use transition is likely to be limited.

While dual use remained persistent, potentially fueling concerns that continued nicotine addiction through ENDS use may make cigarette cessation more difficult, there is evidence of increased harm reduction as well, with an increased fraction of adults who dual used both products having transitioned to ENDS-only use. Although that fraction remains small (overall, we estimated 20.1% of those who used both products transitioned to ENDS-only use in one year), it exceeded the fraction of participants who transitioned from cigarette-only to non-current use (7.1% overall). However, we cannot attribute the increased cigarette cessation from dual use to participants’ use of ENDS; the pattern may be a result of demographic differences between dual and cigarette users or of ENDS use being an indicator of trying to quit. Additionally, our results continue to highlight the importance of understanding the different patterns among different stages of adulthood. Adoption of ENDS among those who currently or previously used cigarettes was much greater among those ages 18–24 (15.3%) than among those ages 25–34 (6.8%) and was nearly negligible among older adults (2.3%) in 2019–21.

There were multiple major public health events and policy changes during the 2019–21 period that could have affected public perception and behaviors around cigarette and ENDS use. Following sharply increased rates of ENDS use among youth in 2018, the US Surgeon General declared youth ENDS use to be an “epidemic”^33^ Less than a year later, the outbreak of vaping-related lung injuries began in August 2019 and continued through the end of the year.^8^ Although the injuries are now thought to largely be associated with the use of vitamin E acetate as a diluent for tetrahydrocannabinol (THC) containing liquids, early CDC reports suggested nicotine vaping as the cause.^8, 34^ The consequent emphasis on dangers of vaping in US news resulted in more negative public perceptions of ENDS use and more of the public discourse focusing on their potential dangers to youth,^35–39^ in contrast to countries like the UK, where there has been more emphasis on ENDS’ potential benefits for adults.^40, 41^ Additionally, in the US, many states and localities enacted restrictions or bans on flavored ENDS sales in 2020 and 2021.^9^

Early 2020 marked the start of the COVID-19 pandemic in the US, as SARS-CoV-2 spread rapidly across the country. Cigarette smoking was quickly found to be a risk factor for severe COVID-19 outcomes,^42–44^ and many studies linked greater perceived risk from COVID-19 to increased motivation to quit smoking.^45–47^ At the same time, elevated stress, particularly during lockdown or similarly restrictive periods, may have led to increased nicotine use as a coping mechanism.^48^ Ultimately, COVID-19 appears to have resulted in a combination of conflicting pressures for and against tobacco and nicotine product use.^49^

Given the period’s many public health events and policy changes, it is perhaps surprising that we do not see more changes in transition patterns for adults who use cigarettes only or ENDS only between 2017– 19 and 2019–21. We did find a small increase in the ENDS-only to non-current use rate, but we found no change in the initiation rate of ENDS among never established users of cigarettes or ENDS nor in the ENDS cessation rate among those using both cigarettes and ENDS. Because the 2019 PATH data collection period encompassed December 2018 to November 2019, the changes we previously found between 2015–17 and 2017–19 may already reflect some of the changes discussed above. Also, because of the local nature of flavor restrictions, the impacts of those restrictions may not be observed at the national scale. Further, any changes in perceptions and behavior may have been short-lived, e.g., if lung-injury-related hesitance to use ENDS in 2019 or COVID-related motivation to quit smoking in early 2020 became irrelevant by the time PATH collected data in 2021. Or, the lack of change may represent the disruption of existing trends, e.g., if more adults who smoke cigarettes would have taken up ENDS in the absence of the lung injury outbreak and flavor restrictions. Because we cannot know what would have happened without these events, we cannot be certain about the extent to which any individual public health event or policy change was associated with the observed transition changes.

The strengths of our analysis include the high-quality, nationally representative data collected by PATH and our multistate transition analysis allowing us to analyze the population-level changes in the transition rates that underly observed changes in transition probabilities over time. Additionally, our results continue to highlight the importance of understanding the different patterns of nicotine product use at different stages of adulthood, given the very different rates of adoption and transition between product use categories between younger and older adults. One limitation of this study is that we did not incorporate data from the Adult Telephone Survey conducted in 2020; we chose to omit these data for this analysis because of the sample size and change in data collection methods. Additionally, our work is limited by our focus only on adults and by a reduced set of age categories compared to previous work.^7^ These changes were made so that the subgroups that had sufficient sizes to support inferences about all the transitions of interest. We plan to analyze youth transitions in separate work with models using a set of product use categories and transitions tailored to the youth population. Finally, our work does not account for the use of other tobacco products, such as cigars, cigarillos, or oral nicotine pouches.

As we discussed above, although many of the changes identified in this analysis appear to be positive from the point of view of reducing cigarette use, the overall public health implications are likely minimal. The US Food and Drug Administration (FDA) is currently determining which ENDS should be authorized and potentially marketed as reduced risk products. Their primary regulatory challenge is to make decisions that encourage cigarette cessation or reduced cigarette use among adults who use cigarettes, while limiting ENDS use among youth and young adults who would never have initiated tobacco use in the absence of ENDS.^18^ While some tobacco-flavored ENDS have received FDA marketing authorization, it is not clear if or when other flavored ENDS could be authorized. In the meantime, a regulation banning non-tobacco characterizing flavors, including menthol, in cigarettes and cigars has been proposed.^50^ It will be important to continue to monitor how transitions in product use continue to change in response to regulatory changes, changes in the marketplace, and future public health events.

## What this paper adds

- It is uncertain how recent changes in the marketplace, as well as events including the lung injury outbreak, COVID-19 pandemic, and ENDS flavor restrictions have impacted real-world product transition patterns.
- Underlying transition hazard rates can explain changes in observed transition probabilities between different patterns of cigarettes and ENDS use.
- We estimated how transition rates and probabilities changed from 2017–19 to 2019–21.
- We found decreased cigarette initiation, increased cessation from ENDS-only use, and increased transitions from dual to ENDS-only use among adults overall. ENDS uptake among cigarette-only users was highest among young adults.
- Dual use remained uncommon (1.9% among adults overall) and ENDS uptake among adult cigarette users remained low (<5% overall), so the population public health impact of the increase in the dual to ENDS-only use transition is likely to be limited.

## Acknowledgments

This project was funded through National Cancer Institute (NCI) and Food and Drug Administration (FDA) grant U54CA229974. The opinions expressed in this article are the authors’ own and do not reflect the views of the National Institutes of Health, the Department of Health and Human Services, or the United States government.

## Contributors

Conceptualisation and methodology: AFB and RMe; data curation: JJ and EJ-M; analysis and original draft preparation: AFB; review and editing: AFB, JJ, EJ-M, SRL, TRH, ASF, JT, RMi, DTL and RMe; funding acquisition: RMe and DTL; guarantor: AFB.

## Data availability statement

Most data are available in a public, open access repository. Other data may be obtained from a third party and are not publicly available. Public Use Files from the Population Assessment of Tobacco and Health Study are available for download from an open access repository (https://doi.org/10.3886/ICPSR36498.v18). Restricted-use files (https://doi.org/10.3886/ICPSR36231.v37) require a restricted data use agreement. Conditions of use are available on the aforementioned websites.

