## Supplemental Material for "Changing patterns of cigarette and ENDS transitions in the USA: a multistate transition analysis of adults in the PATH Study in 2017–2019 vs 2019–2021"

**Multistate transition modeling**

This technical appendix is reproduced in part from the supplementary material of Brouwer et al [6] but includes updated notation and technical details. Additional details may be found in Jackson [32], and a tutorial for working with the weighted multistate transition model (wmsm) code is available at <https://tcors.umich.edu/Resources_Research.php> along with the code itself.

A multistate transition model is a continuous-time, finite-state stochastic process with the Markov assumption that transition rates depend only on the current state and not on past states or transition history. We denote the state of the process at time $t$ as $S\left( t \right)$. We denote the probability that an individual is in state $j$ after an amount of $\Delta t$ since they were observed in state $i$ as

$$\begin{aligned} P_{ij}(t,t+\Delta t)=Pr\left[ S\left( t+\Delta t \right)=j | S\left( t \right)=i \right]. \#\left( 1 \right) \end{aligned}$$

In general, the transition probabilities can depend on the observation time $t$ in addition to the time span $\Delta t$, and in this analysis we consider discrete time periods over which we assume that that the model is homogeneous in time, i.e., $P_{ij}\left( t,t+\Delta t \right)= P_{ij}(0,\Delta t)$. Thus, we drop the dependence on $t$ moving forward and write $P_{ij}(\Delta t)$. We then define the hazard rate of the transition from state $i$ to state$j$, for $i\neq j$, as

$$\begin{aligned} q_{ij}=\lim_{\Delta t\to0} \frac{1}{\Delta t}Pr\left[ S\left( \Delta t \right)=j | S\left( 0 \right)=i \right]. \#\left( 2 \right) \end{aligned}$$

The transition hazard rates form a matrix $Q = \left[ q_{ij} \right],$ where the diagonal entries are given by $q_{ii}=-\sum_{j\neq i} q_{ij}$. The transition probability matrix $P\left( \Delta t \right)=\left[ P_{ij}\left( \Delta t \right) \right]$ is a function of the transition hazard rates and may be calculated as the matrix exponential of $\Delta t\cdot Q$, that is

$\begin{aligned} P\left( \Delta t \right)=\text{expm}\left( \Delta t\cdot Q \right).\#\left( 3 \right) \end{aligned}$

Transition probabilities can be estimate for any value of $\Delta t$, assuming that the transition hazards $q_{ij}$ do not change over that time period (assumption of homogeneity).

The values of the transition hazard rates $q_{ij}$of multistate transition model are estimated by maximizing a statistical likelihood $L$ given a set of observed states and times by comparing the observed states to the probabilities in $P\left( \Delta t \right)$ as a function of the transition hazard matrix $Q$. Specifically, consider a set of individuals $m = 1, \ldots,N$ and their observed states $s_{m,t_{m,k}}$at times $t_{m,k}$, where $k$ is the index of individual $m$’s $K_{m}$observations in the data. Denote the data as $s=\{s_{m,t_{m,k}}\}$. We assume the individuals are independent, and thus we multiply all the modeled probabilities of the observed transitions:

$$\begin{aligned} L\left( Q|s \right)=\prod_{m=1}^{N} \prod_{k=1}^{K_{m}-1} P_{s_{m,t_{m,k}},s_{m,t_{m,k+1}}}(t_{m,k+1}-t_{m,k}).\#\left( 4 \right) \end{aligned}$$

where $P_{i,j}$ is a function of $Q$ as in Eqn (3).

Participant weights $W_{m}$can be incorporated into a weighted likelihood $L^{*}$. Although it is not strictly necessary to normalize the weights to the population size, $w_{m}=N\cdot W_{m}/\sum_{v} W_{v}$, it is convenient to do so because the resulting likelihood will correspond to the unweighted likelihood when all participants have equal weight. The weighted likelihood is given by

$$\begin{aligned} L^{*}\left( Q|s \right)=\prod_{m=1}^{N} \prod_{k=1}^{K_{m}-1} \left( P_{s_{m,t_{m,k}},s_{m,t_{m,k+1}}}(t_{m,k+1}-t_{m,k}) \right)^{w_{m}}.\#\left( 5 \right) \end{aligned}$$

Following the msm package [32], we assume that estimated transition hazard rates $\hat{q}_{ij}$ are normally distributed on the log-scale, that is

$$\begin{aligned} \log\hat{q}_{ij}\sim N\left( \mu=\text{mean}\left( \log\hat{q}_{ij} \right),\sigma^{2}=V\left( \log\hat{q}_{ij} \right) \right),\#\left( 6 \right) \end{aligned}$$

where $V$ denotes the estimated variance.

We estimated weighted point estimates for the log transition hazard rates $\log\hat{q}_{ij}$ by minimizing ${-logL}^{*}\left( Q|s \right)$as a function of the transition hazard rates. (This approach is equivalent to maximum likelihood estimation).

Variance estimates $V\left( \log\hat{q}_{ij} \right)$are calculated using replicate weights $w_{m}^{r}$. Replicate weights are a way to account for complex survey design aspects, such as strata and primary sampling units. PATH uses a variant of balanced repeated replication called Fay’s method to calculate 100 replicate weights. We calculate $\log\hat{q}_{ij}^{r}$ for each $r$ as above. Then, we calculate the variance of $\log\hat{q}_{ij}^{r}$ as

$$\begin{aligned} V\left( \log\hat{q}_{ij} \right)=c\sum_{r=1}^{100} \left( \log\hat{q}_{ij}^{r}-\log\hat{q}_{ij} \right)^{2}\#\left( 7 \right) \end{aligned}$$

where $c=1/(100\left( 1-0.3 \right)^{2})$ as specified by PATH.

Given the mean and variance of two log transition hazard rates, we can test whether two transition hazard rates are statistically significantly different in the following way. Two-sided p-values are calculated as

$$\begin{aligned} 2\left( 1-\Phi\left( x=\left| \hat{\theta}_{1}-\hat{\theta}_{2} \right|; \mu=0, \sigma^{2}=V\left( \hat{\theta}_{1} \right)+V\left( \hat{\theta}_{2} \right) \right) \right), \#\left( 8 \right) \end{aligned}$$

where $\Phi(x;\mu,\sigma^{2})$ is the cumulative normal distribution with mean $\mu$ and variance $\sigma^{2}$, $\hat{\theta}_{1}$ and $\hat{\theta}_{2}$ are estimated log transition hazard rates, and $V\left( \hat{\theta}_{1} \right)$ and $V\left( \hat{\theta}_{2} \right)$are the corresponding variances of the estimated log transition hazard rates.

|  | Adults Waves 2-4 | | Adults Waves 4-5 | | Adults Waves 5-6 | |
| --- | --- | --- | --- | --- | --- | --- |
|  | % | N | % | N | % | N |
| Total | 100 | 24,242 | 100 | 28,061 | 100 | 24,571 |
| Gender |  |  |  |  |  |  |
| Female | 52.1 | 12,591 | 52.0 | 13,522 | 52.0 | 13,075 |
| Male | 47.9 | 11,651 | 48.0 | 14,539 | 48.0 | 11,676 |
| Race/ethnicity |  |  |  |  |  |  |
| Non-Hispanic White | 64.3 | 13,991 | 63.7 | 15,840 | 63.2 | 13,733 |
| Non-Hispanic Black | 15.3 | 4,544 | 15.5 | 5,505 | 15.9 | 5,080 |
| Hispanic | 11.1 | 3,543 | 11.0 | 4,171 | 11.0 | 3,633 |
| Non-Hispanic Other/Unknown | 9.3 | 2,164 | 9.8 | 2,545 | 9.9 | 2,305 |
| Age (years) |  |  |  |  |  |  |
| 18-24 | 14.3 | 7,929 | 12.2 | 8,891 | 12.3 | 8,186 |
| 25-34 | 17.5 | 4,581 | 17.9 | 5,794 | 17.5 | 5,283 |
| 35-90 | 68.2 | 11,732 | 69.8 | 13,376 | 70.2 | 11,282 |
| Tobacco & ENDS use state |  |  |  |  |  |  |
| Never use | 58.8 | 11,968 | 57.9 | 13,894 | 57.7 | 13,023 |
| Non-current use | 21.8 | 4,079 | 23.1 | 5,323 | 23.7 | 4,829 |
| Cigarette-only use | 16.4 | 6,875 | 16.0 | 7,365 | 14.2 | 4,992 |
| ENDS-only use | 1.3 | 564 | 1.5 | 758 | 2.5 | 1,158 |
| Dual cigarette/ENDS user | 1.7 | 756 | 1.5 | 721 | 1.9 | 749 |

***Table S1****: Characteristics of adults in the Population Assessment of Tobacco and Health (PATH) study in 2015–17 (Waves 2–4), 2017–19 (Waves 4–5), and 2019–21 (Waves 5–6), given as weighted percentages (%) and numbers (N).*

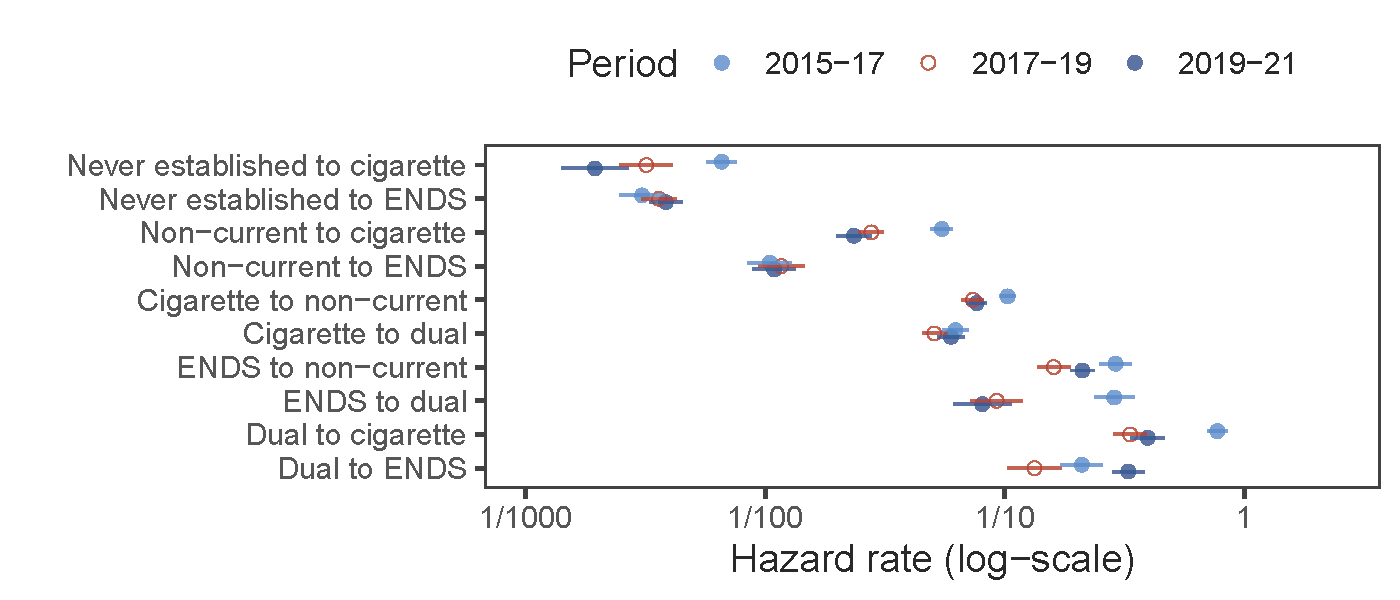

**Figure S1**: *Transition hazard rates among adults in 2015–17 (Waves 2–4), 2017–19 (Waves 4–5), and 2019–21 (Waves 5–6).*

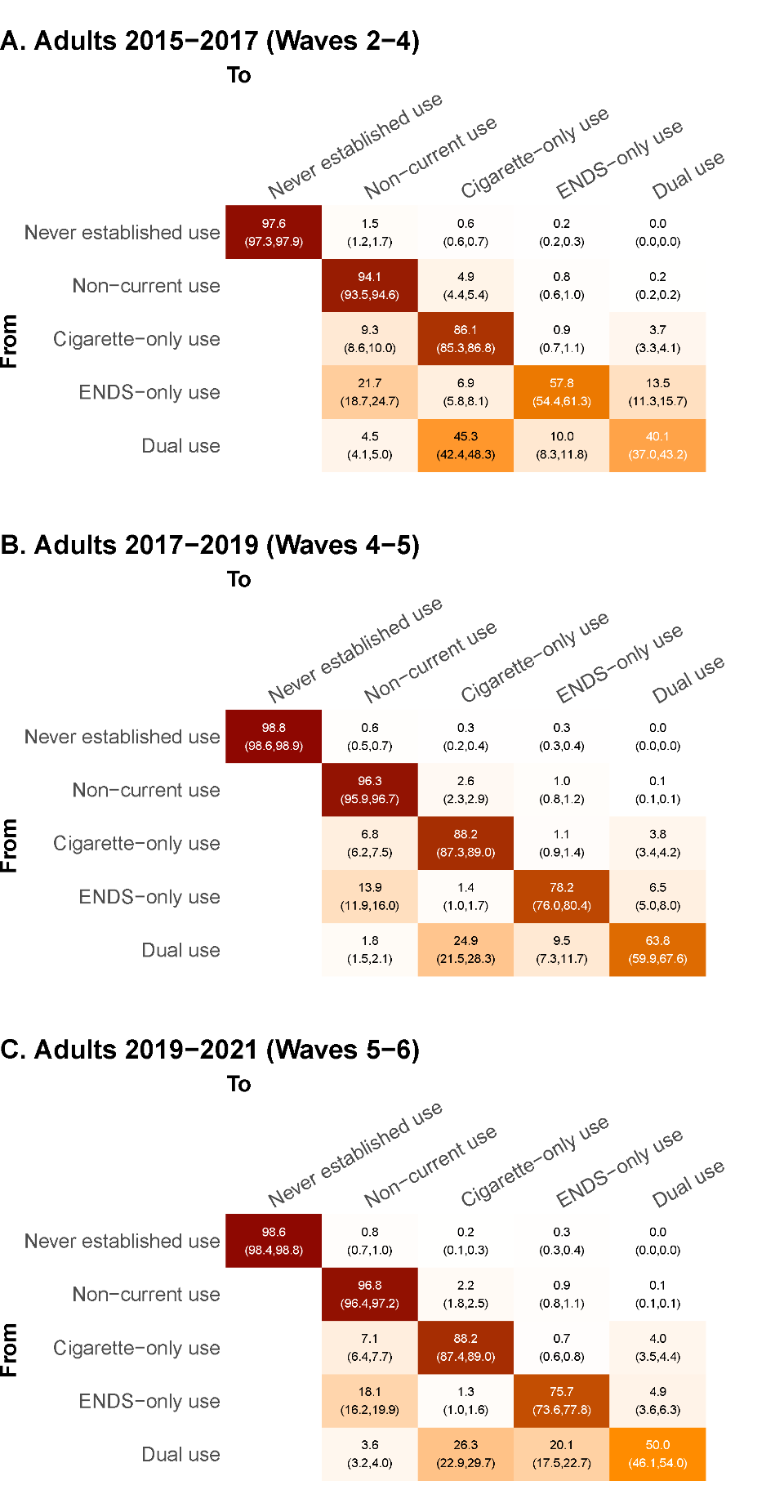

**Figure S2.** *One-year transition probabilities and 95% confidence intervals for adults in A. 2015–17 (Waves 2–4), B. 2017–19 (Waves 4–5), and C. 2019–21 (Waves 5–6).*

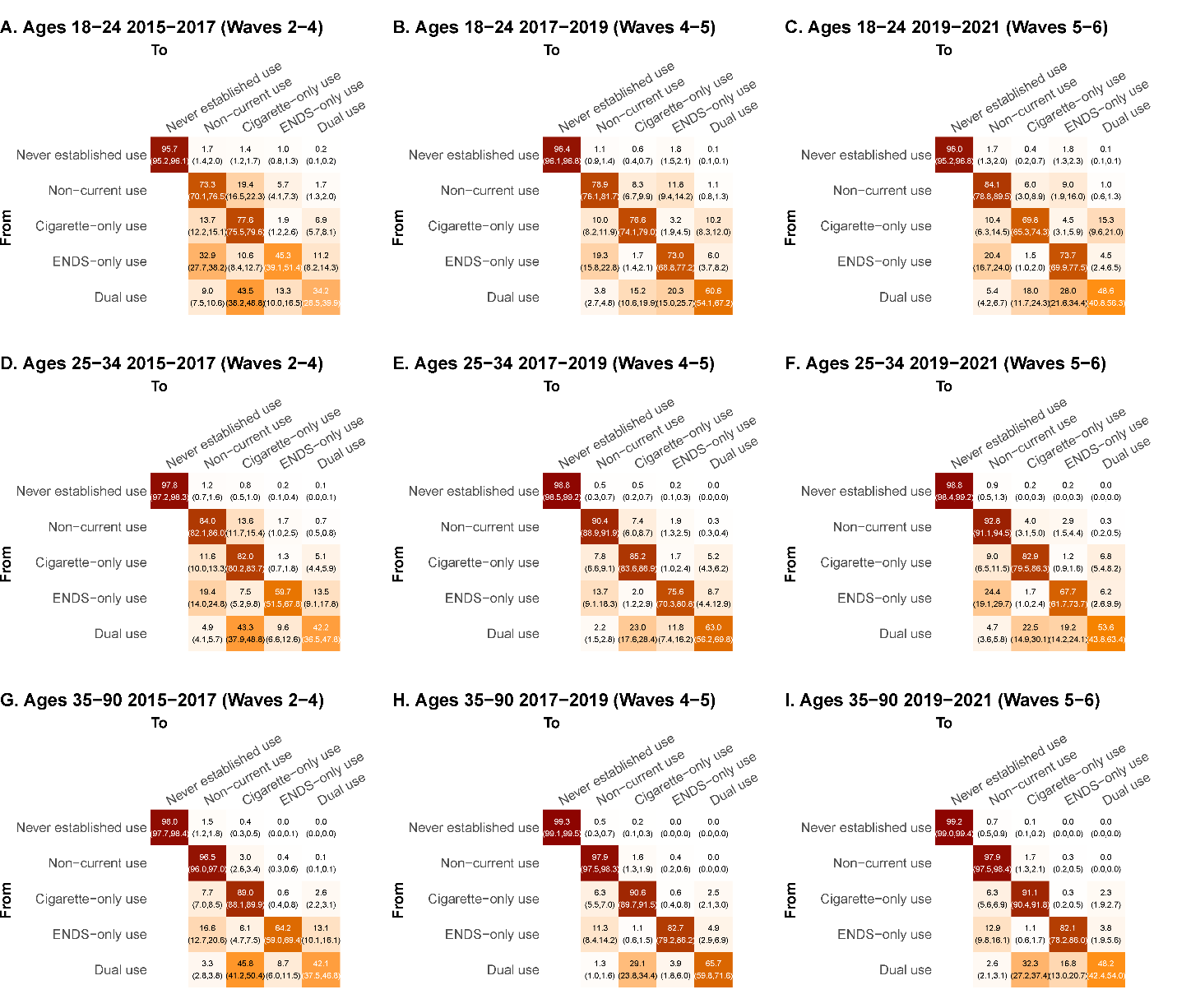

**Figure S3***. One-year transition probabilities and 95% confidence intervals for adults ages 18–24 (A–C), ages 25–34 (D–F), and ages 35–90 (G–I). Each of the three plots compares periods 2015–17 (Waves 2–4), 2017–19 (Waves 4–5), and 2019–21 (Waves 5–6).*

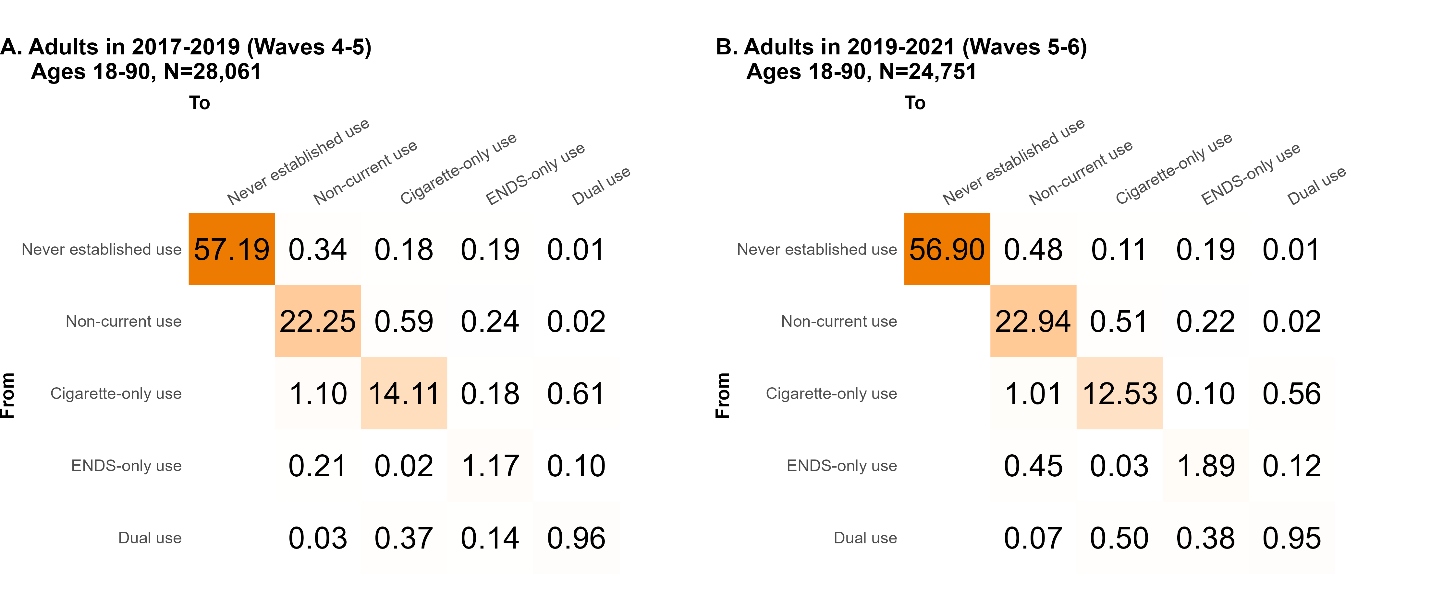

**Figure S4**: *One-year transition probabilities weighted by the prevalence of each use state in that time period given in Table S1, so that all transition probabilities add up to 100% and each row sums to the prevalence of that use type, for adults in A. 2017–19 (Waves 4–5), and B. 2019–21 (Waves 5–6).*
